# Home-based management of COVID-19 by identification of low-risk features

**DOI:** 10.1101/2021.01.25.21249684

**Authors:** Fernando Cabanillas, Javier Morales, José G. Conde, Jorge Bertrán-Pasarell, Ricardo Fernández, Yaimara Hernandez-Silva, Idalia Liboy

## Abstract

**Background:** Covid-19 is a triphasic disorder characterized by a viral phase lasting 7-10 days from first onset of symptoms. In approximately 20% it is followed by a second stage heralded by elevation of pro-inflammatory markers such as ferritin, IL-6, CRP, LDH and D-dimers. We hypothesized that those with few abnormalities would have a low risk for progression to respiratory insufficiency and could be monitored at home without treatment.

**Methods:** Inclusion criteria included age >21, O_2_ saturation >90%. To be observed without treatment patients could not have >1 of the following: CRP > 10 mg/dL, high LDH, ferritin > 500 ng/ml, D-dimer > 1 mg/L, IL-6 > 10 pg/ml, absolute lymphocyte count <1,000, O_2_ sat <94%, or CT chest evidence of pneumonia. Primary endpoint: progression to respiratory failure. Secondary endpoint: 28-day survival.

**Results:** Of 208 entered, 132 were monitored without therapy. None progressed to respiratory failure or died.

**Conclusions:** We have shown that our approach can identify cases who can safely be observed without treatment, thus avoiding expensive, potentially toxic therapies, and circumventing unnecessary, costly hospitalizations. These results support our hypothesis that after applying our criteria, 64% of Covid-19 cases can be monitored as outpatients without therapy.

## Introduction

COVID-19 is a triphasic disorder first typified by an infectious or viral phase that lasts from the first onset of symptoms until 7-10 days later. This is followed by a second phase considered as the inflammatory stage, characterized initially by the appearance of lung infiltrates which is followed in some cases by hypoxemia [1,2]. This second phase is usually heralded by an elevation of serologic inflammatory markers such as C-reactive protein (CRP), ferritin, Interleukin-6 (IL-6), and LDH, as well as D-dimers [3-5]. In a smaller subset of cases this is followed by a third phase characterized by hyperinflammation, leading to the cytokine release syndrome or cytokine storm, that causes Acute Respiratory Distress Syndrome (ARDS) [1,2]. Most COVID-19 deaths are caused by this complication.

Approximately 80% of patients never proceed to the second phase. They are spontaneously cured after the first stage [3,6]. This proportion is a crude estimate that can vary according to several prognostic factors [3-5]. Currently there is no reliable and objective method to accurately predict the 80% that are cured spontaneously without any treatment, vis-à-vis those who develop severe illness. There is consensus that those presenting with severe illness characterized by hypoxemia, should be managed in the inpatient setting, but there are no rigorous, objective criteria to decide when non-hypoxemic patients with mild to moderate illness should be hospitalized instead of monitoring them at home. Traditionally, they are admitted to the hospital if they present with mild to moderate illness without hypoxia but with evidence of pneumonia or dyspnea.

New drugs have recently become available for treatment of mild to moderate COVID-19 infections. These include Remdesivir, and monoclonal antibodies such as bamlanivimab, casirivimab and imdevimab. These new agents have the potential of reducing the number of hospitalizations if applied early during the infectious process, however their cost can be quite high. In addition, all these agents have potential toxicity. The overuse of these drugs could be avoided if we have a method to identify low-risk cases who do not require any therapy and who could be safely monitored outside of the hospital setting, hence further reducing the financial costs.

Furthermore, excessive hospitalizations can overwhelm and overload healthcare systems. A method to identify cases that can be safely managed on an ambulatory setting could relieve this burden. Finally, knowledge regarding the factors that predict the likelihood of disease progression could help physicians decide who can be managed safely at primary care facilities and who needs to be transferred to a tertiary care center.

We hereby report on the results of the application of the risk-stratification stage of this phase II clinical trial designed with two goals in mind:

1. To identify early during their illness low-risk patients who can be safely monitored at home without treatment.
2. To identify patients with COVID-19 at high-risk of progressing to hypoxemic respiratory failure, so they can be treated prophylactically with steroids early during their clinical course to prevent them from developing ARDS or cytokine release syndrome.

We prospectively classified COVID-19 patients by ranking them into high and low-risk, according to blood-based biomarkers of inflammation, as well as other clinical features. We then analyzed the clinical outcome of those considered as low-risk. The current manuscript exclusively describes the outcomes of low-risk cases. The results related to the management of high-risk cases will be the subject of a separate report.

## Materials and Methods

### Investigational plan

The study was registered in clinicaltrials.gov as NCT04355247 and registered as such with the local Institutional Review Board at Auxilio Mutuo Hospital which approved it. Informed consent was obtained from each patient prior to recruitment.

The original plan was to enter a total of 100 patients with the expectation that at least 20 would-be high-risk cases eligible for therapy with methylprednisolone and 80 would be low-risk, eligible for monitoring without therapy. This expectation was based on the available literature data [3,6] which describes a 20% chance for patients with COVID-19 to develop severe disease associated with respiratory failure. We later decided to expand this pilot study to include at least 200 patients.

When the CALL score method to predict prognosis was published ^2^, we amended the protocol to apply this method with the purpose of allowing us to more precisely predict the expected number of cases that would develop respiratory failure. Using this prediction, we could then calculate the expected number of cases and compare it with our therapeutic results using our risk-assessment method. This CALL Score method considers the presence of comorbidities, age, LDH level and lymphopenia to assign a prognostic score. The higher the score, the worse the prognosis. A nomogram which is part of the CALL score, was used to predict the risk of progression to respiratory failure [8].

### Eligibility

Eligible patients for this protocol had to be over 21 years old with a diagnosis of COVID-19 established by means of either the PCR molecular test (97% of enrolled cases) or with the rapid serologic test in the context of typical symptoms and/or ground glass infiltrates in the chest CT (3% of cases). There was no top age limit for entry. Excluded from entry were those who already were in acute respiratory failure defined as oxygen saturation <91%. Other exclusion criteria included subjects who were oxygen dependent, or who had long standing history of severe COPD. Anyone receiving tocilizumab, convalescent plasma therapy or prednisone 20 mg daily or equivalent were also excluded.

### Definition of low-risk cases

For patients to be classified as low-risk, they could have only one or none of the following abnormalities between days 7-10 after their first symptom: IL-6 ≥ 10 pg/ml, Ferritin> 500 ng/ml, D-dimer > 1 mg/L (1,000 ng/ml), CRP > 10 mg/dL (100 mg/L), LDH above normal, lymphopenia (absolute lymphocyte count <1,000), oxygen saturation between 91-94%, or CT chest with evidence of ground glass infiltrates. These markers were selected based on published data which have shown a strong correlation with a poor outcome [10 -15]. We also analyzed the symptomatology at presentation to determine if low-risk patients could be identified by the number of symptoms at diagnosis or by the type of presenting symptoms.

### Management

Patients classified as low-risk were monitored at home without treatment. They were asked to check their oxygen saturation three times per day by means of pulse oximetry and to report any value less than 94% during the first two weeks after entry. They were also instructed to immediately report any unexpected change in their clinical condition. Patients were also called daily for 28 days to inquire about their condition.

### Statistical analysis

Medians and interquartile ranges (IQR’s) were used to describe distributions of continuous variables, and proportions to describe distributions of categorical variables. IQR’s are reported as lower and upper limits of the IQR (i.e. first and third quartiles of distributions). This format not only provides information on the IQR, but also indicate location of the central 50% of distributions relative to the median. The Wilcoxon-Mann-Whitney test was used for testing hypotheses about the difference between distributions of continuous variables, and the chi-square test for categorical variables [16].

## Results

We initially enrolled 213 patients, 5 of which were not evaluable because consent was withdrawn (N=3), lost to follow up after suicidal attempt (N=1), PCR for Covid-19 negative (N=1). Of the remaining 208 patients, 132 (63.5%) fulfilled criteria for low-risk, hence they met conditions for monitoring at home without therapy, while the remaining 76 were classified as high-risk and treated with methylprednisolone.

The median follow-up time for these 132 low-risk cases was 84 days (range 31-263) for a total of 10,956 person-days of follow up with interquartile range of 44. At the time of diagnosis, 97 (73.4%) of the low-risk cases, were polysymptomatic, presenting with 2 or more of the following symptoms: fever, cough, myalgia, diarrhea, anosmia, dyspnea, or headache. Only 15 of the low-risk cases were either asymptomatic (11 cases) or minimally symptomatic (4 cases) with nasal congestion and sore throat, suggesting that low-risk cases, as defined by our criteria, cannot be accurately identified by using the number or types of symptoms at presentation. In addition, there were 48 cases who had a chest CT done at time of diagnosis and of these, 17 presented with typical ground-glass infiltrates.

Table 1 summarizes the baseline characteristics of all 208 cases including low as well as high-risk cases. Male gender was more commonly represented in high-risk patients, but this did not reach statistical significance (p=0.36). High-risk patients were significantly older than low-risk. Similarly, the number of comorbidities was significantly greater in the high-risk cases. The CALL score correlated well with the risk assignment. Most commonly, the low-risk cases corresponded to the favorable CALL score “Class A group”, and only 2% were unfavorable “Class C group”. However, there was a relatively large proportion, 47%, that fell into the intermediate CALL score “Class B group”. As expected, high-risk cases presented more commonly than low-risk cases with CALL score “Class C” as well as with lower oxygen saturation. Regarding symptoms, fever and dyspnea were more commonly seen in high-risk cases.

**Table 1.**

| Feature | Low risk<br>(N=132) | % | High-risk<br>(N=76) | % | P |
| --- | --- | --- | --- | --- | --- |
| Gender |  |  |  |  |  |
| Male | 62 | 47 | 40 | 52.6 | 0.36 |
| Female | 70 | 53 | 36 | 47.3 |  |
| Median age (IQR) | 45 (22) | - | 60 (23) | - | .00001 |
| Median #<br>comorbidities per<br>patient (IQR) | 1 (1) | - | 2 (2) | - | .00001 |
| CALL Score: |  |  |  |  |  |
| 4-6 (Class A) | 64 | 48.5 | 14 | 18.4 | .0001 |
| 7-9 (Class B) | 62 | 47.0 | 36 | 47.3 |  |
| 10-13 (Class C) | 3 | 2.2 | 26 | 34.2 |  |
| Missing information | 3 | 2.2 | 0 | 0 |  |
| O<sup>2</sup> saturation 91-94% | 2 | 1.5 | 23 | 30.2 | .00001 |
| Symptoms: |  |  |  |  |  |
| Fever | 49 | 37.1 | 48 | 63.2 | .0001 |
| Dyspnea | 21 | 15.9 | 29 | 38.2 | .004 |
| Myalgia | 84 | 63.6 | 49 | 64.4 | 1.0 |
| Diarrhea | 55 | 41.7 | 33 | 43.4 | 0.88 |
| Anosmia | 67 | 50.7 | 34 | 44.7 | 0.47 |

There were 3 cases in the low-risk category for which there was missing information because their CBC was reported without a differential count, necessary for the calculation of the CALL score. Since these three cases had no other high-risk criteria, they were assigned to the low-risk group.

The presence of dyspnea and pneumonia are frequently used as criteria to decide on inpatient management. There were 48 of our low-risk patients who had a chest CT done and 17 of these had typical findings of ground-glass infiltrates that would traditionally have required hospitalization. An additional 9 cases had a negative chest CT but complained of dyspnea. Another four had dyspnea without hypoxemia, hence a CT chest was not done. In summary, 30 cases met traditional conditions for admission, yet were managed as outpatients in view of their low-risk features after applying our criteria.

### Clinical Outcomes

None of the 132 low-risk cases developed respiratory failure and none developed any type of complication that required admission to the hospital while under follow-up. None of them died.

Four (1.9%) of the 208 patients developed what appeared to be a reinfection. All four were either asymptomatic or minimally symptomatic at time of first diagnosis. After applying our criteria, all four were classified as low-risk cases. They had been positive by PCR at time of first diagnosis but subsequently turned PCR negative several days later to again become positive at the time they developed new symptoms suggestive of reinfection.

The first case of reinfection is a female in her mid-fifties diagnosed after having been exposed to a patient infected with COVID-19. She was asymptomatic at time of diagnosis. Her first PCR was positive but 7 days later it turned negative. At time of diagnosis her serological studies were negative for IgG and IgM. She remained asymptomatic until 4 months later when she suddenly developed fever, dyspnea, cough, anosmia, and myalgia and her PCR then turned positive. Based on inflammatory markers, including LDH and IL-6, she met criteria for treatment during this second bout. In addition, her oxygen saturation was 93% and CT chest revealed changes compatible with COVID-19 pneumonia. She was treated with methylprednisolone 80 mg daily IV x 5 days and rapidly improved clinically. By day seven, her inflammatory markers also improved.

The second reinfected case is a female in her low fifties who had been exposed to a co-worker who had been diagnosed with COVID-19. She was asymptomatic but on day 7 post exposure, her PCR test was positive. She did not fulfill high-risk criteria, so she was monitored without therapy. Ten days after the positive PCR test, IgM and IgG antibody tests were both negative. Two weeks after the first PCR test, the PCR was repeated and became negative. She remained well until 20 days after the first PCR, when she developed fever, diarrhea, cough, anosmia, and myalgia. Her PCR was repeated and was now found to be positive. A confirmatory PCR was ordered 18 days later and again was positive. Nine days after her second infection, antibody tests for IgM and IgG both turned positive. This time, based on inflammatory markers, she met criteria for treatment with methylprednisolone and she was started on therapy according to protocol. She responded clinically and her symptoms resolved promptly. However, a D-dimer test was repeated as part of the protocol on day 7 post methylprednisolone, and it abruptly increased from a baseline of 0.38 to 6.36 mg/L. She developed severe coughing spells and this clinical picture brought up the suspicion of a thrombotic complication secondary to COVID-19. CT chest angiogram was done which showed several right sided pulmonary emboli. Anticoagulation with rivaroxaban was initiated and her coughing spells rapidly subsided.

The third case of reinfection was a male in his mid-twenties who developed nasal congestion and a positive PCR test. Eight days after this PCR, his IgG and IgM antibodies were negative, and a follow up PCR test converted to negative. He fulfilled criteria for low-risk disease, so he was observed without therapy. He did well until four months later when he was exposed to several family members who were infected with COVID-19 and shortly after, he developed typical symptoms of COVID-19 consisting of fever, myalgia, anosmia, headache, and diarrhea. One week later his PCR turned positive. His IgG and IgM also turned positive 8 days after this last PCR. By the time we received the PCR result, patient was already recovering from his symptoms, so we decided not to treat him. Since we did not order an earlier confirmatory PCR after the first infection, we cannot completely rule out the possibility that instead of a reinfection, his first PCR could have been a false positive result, although this is unlikely.

The fourth case of reinfection is a male in his mid-thirties who had a routine PCR test ordered on a routine basis at his workplace. He was essentially asymptomatic except for diarrhea which he had first noticed two days earlier. The PCR test was positive. It was repeated one week later and again was positive. Eight days after his first symptom, his IgG and IgM antibodies were negative. Six weeks after his first symptom, his PCR converted to negative. He fulfilled criteria for low-risk disease, so he was observed without therapy. He remained well until six months later when he again developed diarrhea but this time, he also had cough, fever, myalgia, fatigue, and headache. PCR was ordered and it was positive. A confirmatory PCR was obtained four days later which was again positive. On day 8 from his first symptoms of reinfection, IgG was positive and IgM negative. He did not meet criteria for treatment with methylprednisolone and thus was monitored without therapy. He remains well without complications.

## Discussion

This manuscript focuses exclusively on 132 low-risk cases. The remaining 76 cases will be the subject of another report. Our management approach, based on blood-based inflammatory markers as well as other clinical features, had an excellent correlation with clinical outcome. Of 208 patients, 132 or 63.5%, fulfilled criteria for low-risk disease that justified monitoring at home without therapy. None of these low-risk cases developed respiratory failure and none developed any significant complication or required admission to the hospital. Included in this low-risk group as defined by our criteria, were 68 cases with comorbidities, 17 with COVID-19 pneumonia, as well as with other adverse features, including 15 cases older than 65 and 96 with more than two symptoms at presentation. These characteristics would usually portend an unfavorable prognosis according to traditional standards, yet all these patients had an excellent outcome.

The management of mild to moderate COVID-19 infections has traditionally been conservative. Patients with either COVID-19 pneumonia or with dyspnea, are often admitted to the hospital ^1^. Otherwise, treatment has consisted of observation at home provided there is no evidence of hypoxemia. Applying these traditional criteria to decide on hospitalization, 26 of our 48 low-risk patients who had a chest CT, would have been managed as inpatients because 17 had typical findings of ground-glass infiltrates that would traditionally have been hospitalized and an additional 9 cases had a negative chest CT but complained of dyspnea. Another 4 cases (20%) had dyspnea without hypoxemia and hence a chest CT scan was not done. This adds up to a total of 30 patients who traditionally would have been managed as in-patients. By applying our novel criteria to identify low-risk cases, we were able to safely manage all these 30 patients at home without separating them from their families. Furthermore, assuming an average of 10 days of hospitalization per patient, it allowed us to save an average of 300 days of hospitalization. At an average cost of hospitalization of $3,949 per day, this adds up to a total of $1,184,700 US dollars saved. In addition, these 30 patients were prevented from being exposed to nosocomial infections.

Dexamethasone has been utilized to treat hospitalized patients with COVID-19. Those who are ventilator-dependent or who are receiving oxygen have benefitted from that treatment, while those who are oxygen-independent fail to do so and might even be adversely affected since their mortality rate tends to be higher when treated with dexamethasone [9]. It is conceivable that some patients are unnecessarily being treated with this medication, and our proposed method should be able to identify them.

After the first week of illness, 20% of patients diagnosed with COVID-19 might suddenly deteriorate and develop respiratory failure [3,6]. This includes those who initially present with mild symptomatology

. Consequently, being able to prospectively segregate those cases who will have an uncomplicated clinical course from those who are more likely to develop respiratory failure and who could potentially benefit from preemptive immunosuppressive therapy is an important advantage.

Although there were statistically significant differences between low-risk and high-risk cases regarding their clinical presentation and symptoms (Table 1), there was not a single factor or factors in their clinical presentation that could reliably be used to classify them into either the low-risk or high-risk groups. The frequency of symptoms would intuitively be expected to be less in the low-risk cases. However, contrary to our expectations, the majority (73%) of low-risk cases presented with two or more symptoms at diagnosis, indicating that they cannot be identified based on symptomatology alone. The CALL Score is an exceptionally useful method. It assigns a risk score which can be used to determine the probability of a given patient to progress to respiratory failure. It is based on quite simple and straightforward clinical features. Nonetheless, neither the CALL score nor any other system that we are aware of, was able to identify the low-risk cases detected by our novel criteria.

It is interesting that of the four cases that were possibly reinfected, all four presented with mild disease at the time of diagnosis. It is well known that patients with mild COVID-19 infections produce little or no antibodies [17] so it should not be surprising that all our four cases with possible reinfection had mild or no symptomatology at the time of initial diagnosis. Unfortunately, we could not perform molecular analysis of the virus at time of reinfection to determine if a different viral strain was involved.

Our results suggest that by applying our novel criteria, low-risk cases can be reliably monitored at home without treatment, rather than immediately embarking on admission to a hospital or with outpatient treatment. Aside from the positive financial and psychological aspects of home-based management, the ability to identify low-risk cases at diagnosis has several other advantages, including the possibility of circumventing the use of expensive or potentially toxic agents.

Although the data generated by this prospective trial appear convincing, it is important to keep in mind that this is a pilot study and confirmation of these findings in another cohort or in an independent study would be highly desirable.

## Data Availability

No external datasets or supplementary material.

